# A Multicenter, Controlled, Randomized Trial of Home-Applied Dual-Light Photodynamic Therapy in Chronic Periodontitis

**DOI:** 10.1101/2025.03.25.25324596

**Authors:** Paula Tegelberg, Meeri Ojala, Merja Ylipalosaari, Jori Lindroth, Anna-Maria Heikkinen, Hanna Lähteenmäki, Saila Pakarinen, Ismo T. Räisänen, Timo Sorsa, Tommi Pätilä

**Affiliations:** Research Unit of Population Health, Faculty of Medicine, University of Oulu, Oulu, Finland; Institute of Dentistry, University of Eastern Finland, Kuopio, Finland; Faculty of Medicine and Health Technology, University of Tampere, City of Tampere Finland; Department of Oral and Maxillofacial Diseases, Faculty of Medicine, University of Helsinki, and Helsinki; Metropolia University of Applied Sciences, Helsinki, Finland; Department of Congenital Heart Surgery and Organ Transplantation, New Children’s Hospital, University of Helsinki, 00290 Helsinki, Finland

**Keywords:** Periodontitis, Antibacterial photodynamic therapy (aPDT), Dual-light photodynamic therapy, Non-surgical periodontal treatment (NSPT), Photobiomodulation therapy (PBMT)

## Abstract

**Aim:** This multicenter, randomized controlled trial evaluated the efficacy of the Lumoral® dual-light antibacterial photodynamic (aPDT) home care device as an adjunct tool to non-surgical periodontal treatment (NSPT) in chronic periodontitis patients.

**Materials and Methods:** Forty-one Stage I-III periodontitis patients were randomized to receive NSPT, including standardized hygiene instructions and scaling and root planing, or NSPT with adjunctive Lumoral® treatment, applied daily at home. Clinical examinations were conducted at baseline, three months, and six months.

**Results:** Both groups showed significant reductions in Visual Plaque Index (VPI) and Bleeding on Probing (BOP) at three and six months. However, the NSPT + Lumoral® group exhibited significantly greater BOP and VPI reductions at six months (p=0.03, p=0.04) compared to the NSPT group. Incidence of Probing Pocket Depth (PPD) ≥ 4 mm reduced significantly in the NSPT + Lumoral® group at both follow-ups (p=0.0019, p=0.0043) but not in the NSPT group. Periodontal epithelial surface area (PESA) and periodontal inflamed surface area (PISA) significantly decreased in the NSPT + Lumoral® group (p<0.01), while no significant changes were observed in the control group.

**Conclusion:** Regular adjunctive use of Lumoral® after NSPT resulted in improved clinical periodontitis treatment outcomes.

## 1. Introduction

Periodontitis is a chronic inflammatory disease affecting the attachment and supporting structures of the teeth. Its infection-induced pathogenesis is driven primarily by subgingival dysbiotic microbiota and the host immune response, ultimately leading to the irreversible destruction of soft and hard periodontal tissues [1]. Antibacterial photodynamic therapy (aPDT) and antibacterial blue light (aBL) have emerged as promising approaches to reduce the burden of dental biofilms [2–5]. While aPDT has demonstrated antibacterial efficacy, its effectiveness in periodontitis management has been inconsistent [6]. One evident explanation is that most clinical studies have applied aPDT only once or twice, which may not be sufficient for sustained therapeutic benefits [7,8]. In contrast, studies incorporating repeated aPDT treatments have reported more favorable outcomes [9–11], suggesting that treatment frequency, indeed, may play a critical role in clinical outcome.

Dual-light therapy, which combines aPDT and aBL, exerts significant antibacterial activity in dysbiotic biofilms and has been associated with reduced dental plaque formation [9,12–14]. Lumoral® is a CE-marked medical device (Koite Health Ltd., Espoo, Finland) delivering simultaneous 405 nm aBL and 810 nm near-infrared (NIR) LED light through a mouthguard-based system. When used with Lumorinse®, an indocyanine green (ICG) mouth rinse (Koite Health Ltd., Espoo, Finland), the treatment enables targeted antibacterial action due to a selective adherence of ICG to dental plaque and bacteria [12,13]. In regular home use, dual-light therapy with Lumoral® has been shown to reduce inflammation and inflammatory markers in peri-implant disease and has demonstrated benefits in supporting oral hygiene, and it has proven to be safe to use [9,15,16].

This clinical study was designed to evaluate the effectiveness of home-applied dual-light aPDT in improving periodontal disease symptoms and clinical outcomes. We hypothesized that when used in conjunction with non-surgical periodontal treatment (NSPT), regular antimicrobial therapy would reduce the dysbiotic microbial burden on gingival tissue. Additionally, the near-infrared component of the dual-light therapy may provide a photobiomodulation therapy (PBMT) effect, further supporting and promoting periodontal tissue healing.

This study is the first multi-center clinical trial investigating continuous, repetitive home-based aPDT treatment. The primary outcome measure was a reduction in gingival inflammation, assessed by bleeding on probing (BOP). (ClinicalTrials.gov Identifier: NCT05425784).

## 2. Materials and Methods

### Study design

This study is a multicenter, randomized clinical trial designed to evaluate the efficacy of regular home use of the dual-light aPDT in patients diagnosed with periodontitis. The study protocol was approved by the Regional Medical Research Ethics Committee of the Wellbeing Services of North Ostrobotnia, Finland (EETTMK/27/2022). The study was conducted in accordance with the ethical principles of the Declaration of Helsinki, the good clinical practice (GCP) ISO 14155, and Medical Device Regulation 2017/745. Three dental clinics in Northern Finland participated in the study. All participants provided written informed consent. Patients were randomised to receive either NSPT, which included standardised hygiene instructions, an electric toothbrush and scaling and root planing (NSPT group), or NSPT with adjunct Lumoral® treatment (NSPT + Lumoral® group).

### Sample size

This report includes 41 consecutive patients from three different sites in Northern Finland, of which one patient was a screening failure and was subsequently replaced. For analysis, there were 20 patients in the NSPT + Lumoral® group and 20 in the NSPT group. All subjects over 18 years of age were eligible for enrolment according to the following inclusion and exclusion criteria.

### Eligibility criteria for study participants

The study included participants with Stage I-III periodontitis based on the 2017 World Workshop on the Classification of Periodontal and Peri-Implant Diseases and Conditions [17]. Subjects were referred for professional dental treatment through each individual’s oral health care appointment system with an already diagnosed or suspected case of periodontitis.

#### Inclusion criteria

- Periodontal disease stage I-III, based on the 2017 World Workshop on the Classification of Periodontal and Peri-Implant Diseases and Conditions with at least 2 mm interdental CAL in the site of greatest loss,
- Age of 18-85 years,
- Presence of ≥20 teeth, including implants,
- Agreement to participate in the study and to sign a written consent form, and
- Ability to cooperate with the treatment protocol and avoid any other oral hygiene measures outside of the study protocol.

#### Exclusion criteria

- Presence of major physical limitation or restriction that might prohibit the hygiene procedures used in the study protocol,
- Removable major prosthesis or major orthodontic appliance,
- Pregnancy or lactation,
- Use of antibiotics within 2 weeks prior to the study, and
- A need for immediate antimicrobial treatment for periodontitis.

### Randomization

A block randomization method using sealed envelopes was employed to ensure an equal distribution of participants between the control and treatment groups. Participants were randomly assigned in a 1:1 ratio to either the NSPT + Lumoral® group or the NSPT group, using blocks of ten to maintain balance. The sealed, sequentially numbered envelopes were securely stored in the Sponsor’s Research Manager’s office. When an investigator requested randomization, the Research Manager or an authorized staff member opened the next available envelope and informed the investigator of the participant’s assigned group. The investigator immediately recorded the assignment in the participant’s file, while the Research Manager documented the study number in the randomization log.

### Intra-examiner reproducibility

Clinical measurements were performed at three study sites by three experienced examiners (P.T., M.O., and M.Y.). Before the study began, the examiners underwent calibration for probing pocket depth (PPD) and clinical attachment level (CAL) to ensure consistency. Using a manual probe (LM 8-520B, Lääkintämuovi, Finland), they assessed six sites per tooth within one sector for a total of four subjects. Calibration consisted of two measurement sessions conducted no more than 48 hours apart. PPD and CAL values were recorded and compared between sessions, with calibration considered acceptable if at least 85% of the measurements remained consistent within one millimeter across both sessions. The intra-examiner reproducibility was 100% for M.O., 93% for M.Y., and 94% for P.T., confirming the successful calibration of the examiners.

### Clinical procedure

Demographic data were collected at baseline. Clinical intraoral examination including BOP, visible plaque index (VPI), CAL, and PPD were obtained at baseline and at the 3 and 6-month follow-up visits.

After baseline measurements, all patients received standard anti-infective treatment. Patients in the control group received no additional intervention, while patients in the study group received the home-use dual-light aPDT Lumoral® treatment device including the Lumorinse® mouth rinse tablets for six months.

### Anti-infective treatment

Scaling and root planning (SRP) was thoroughly performed, including the removal of biofilm and calculus from all surfaces. The SRP was performed with an ultrasonic instrument NSK Varios 750 with scalper tips G6 and G9 (NSK Dental, Kanuma, Japan) or Acteon Satelec Newtron with 10Z, PFL, or PFR scaler tips (Acteon Group, Norwich, UK) with water cooling, and/or hand instruments (LM Gracey curettes, LM-Dental Instruments, Parainen, Finland). After the SRP, pasta cleaning was performed with a relative dentin abrasivity of 250 (Topdent, Shanghai, China), and fluoride varnish was applied if needed.

In addition to the anti-infective treatment, standard oral hygiene instructions, including the use of an electric toothbrush, interdental brush, silicone interdental toothpick, and dental floss, were given to all participants. All patients were given a new electric toothbrush to ensure consistent access to good oral self-care standards (Jordan AS, Oslo, Norway).

### Clinical measurements

All clinical measurements were recorded using a ball pointed periodontal probe with 2 mm grading (LM 8-520B, Lääkintämuovi, Finland) with a maximum force of 0.25 N for six sites (mesiobuccal, buccal, distobuccal, mesiolingual, lingual, distolingual) of each tooth.

BOP was assessed at six sites per tooth, based on the presence or absence of gingival bleeding within 15 seconds of gentle probing, and reported as the percentage (%) of sites with positive full-mouth findings. Full-mouth VPI was assessed using a six-point dichotomous scoring system as plaque ‘1 present’ or ‘0 absent’. VPI is presented as a percentage (%) of sites with positive findings. PPD was measured in millimeters from the gingival margin to the base of the periodontal pocket. Gum recession was assessed by measuring the distance from the enamel-cementum junction to the gingival margin, while gingival overgrowth was recorded accordingly.

### Periodontal Epithelial Surface Area (PESA)

PESA represents the total surface area of the periodontal pocket epithelium. It is calculated by determining the root surface area affected by clinical attachment loss (CAL) and subtracting the recession surface area (RSA) from the attachment loss surface area (ALSA), measured at six sites per each tooth. This quantification provides an estimate of the epithelial interface exposed in periodontal pockets but does not distinguish between inflamed and healthy epithelium. The PESA values were calculated based on deep periodontal pockets, defined as sites with PPD≥ 4 mm, and assessed from six sites of all teeth.

PESA represents the total epithelial surface area (i.e. periodontal pockets), while PISA quantifies the inflamed surface area by calculating the incidence of bleeding around the gums of a tooth representing each individual periodontal pocket. The health of the gums and the degree of inflammation in the periodontium were evaluated by calculating the total periodontal epithelial surface area (PESA) and the total periodontal inflamed surface area (PISA). This provides an objective measure of periodontal inflammation.

### Periodontal Inflamed Surface Area (PISA)

PISA quantifies the inflamed portion of the periodontal pocket epithelium by incorporating BOP data, measured from six sites per tooth. It was derived by multiplying the PESA by the proportion of sites exhibiting BOP. This measurement expresses the periodontal inflammatory burden in square millimeters, offering a more precise indicator of the systemic inflammatory potential of periodontitis.Both PESA and PISA calculations are facilitated using an Excel-based computational tool, where full-mouth CAL, recession, and BOP data from six sites per tooth are entered [18].

### Dual-light aPDT treatment

The Lumoral® treatment device is a CE-marked antibacterial home device for the treatment and prevention of oral diseases caused by bacteria. It is used in combination with a CE-marked mouth rinse called Lumorinse®. See Figure 1.

**Figure 1.**
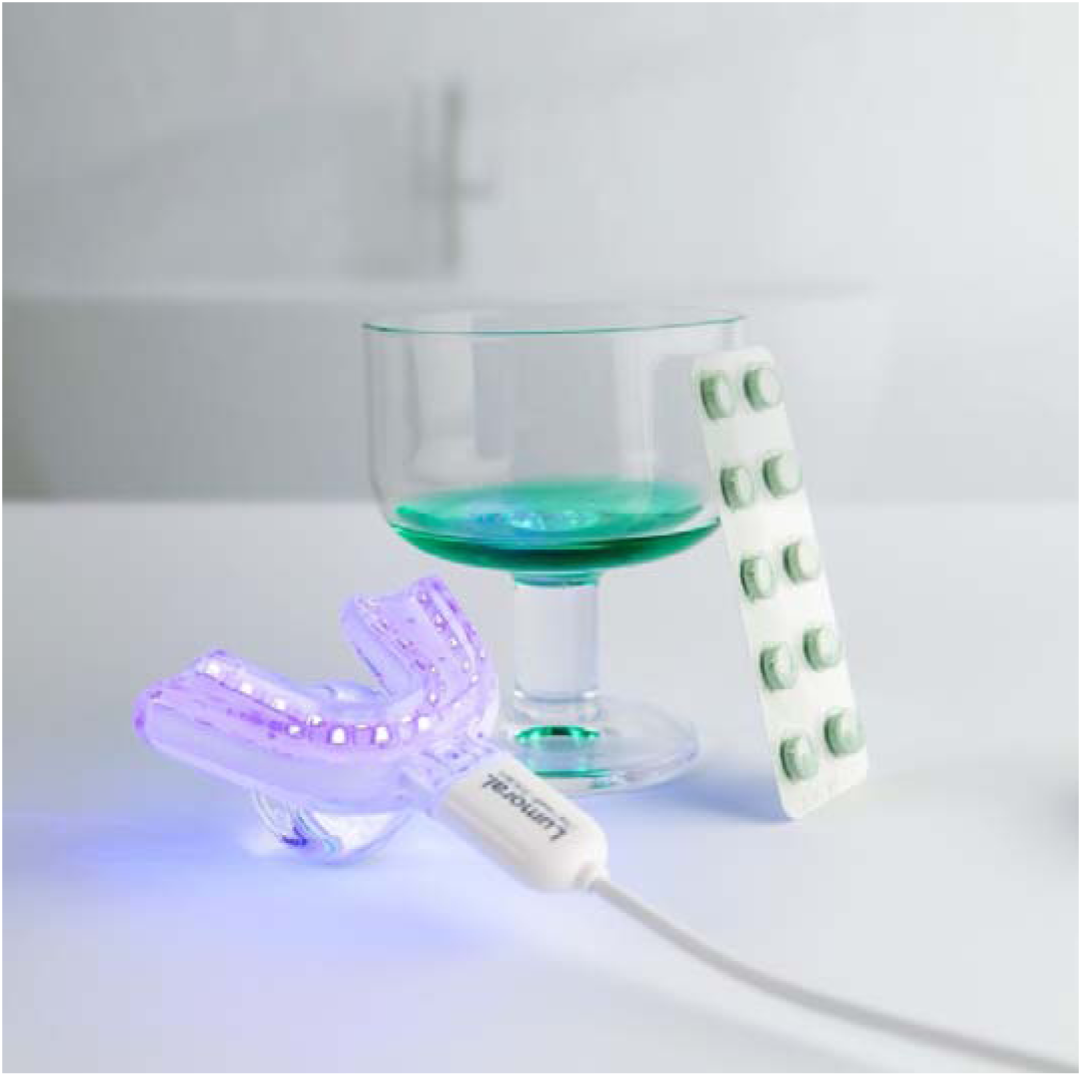
Lumoral device consists of photosensitizer mouthrinse and a light applicator. The mouthrinse is provided in an effervescent tablet form, and it is diluted in water before use. The light applicator is provided in one size, and it has 48 dual light LED components emitting light for both dental arches simultaneously.

Patients received detailed instructions on how to use the Lumoral® device with accompanying Lumorinse® mouth rinse tablets and were instructed to use the Lumoral® device daily, but at least five per week and follow the protocol.

The Lumorinse® mouth rinse is an effervescent tablet with a final ICG concentration of 250 μg/mL. The mouth rinse is swirled for 60 seconds to allow the ICG to adhere to the plaque and spit out. When adhered to the plaque, the ICG is activated by the light activator, or Lumoral® treatment device, which provides activating light simultaneously at both the lower and upper dental arches. Lumoral® has 48 LED components that deliver two separate wavelengths described as dual light action. The near-infrared 810 nm light activates the plaque-attached IGC, and the 405 nm blue light delivers the simultaneous aBL-action. The blue light is absorbed by the intrinsic chromophores within the bacteria, mainly porphyrins and flavins, which then release the light energy leading to bacterial elimination. After 10 minutes and 30 J/cm2 exposure, the device switches off automatically. The treatment is used in addition to regular dental hygiene procedures such as toothbrushing and interdental cleaning.

### Compliance and adverse events reporting

Patients in the study group were asked to keep a diary of Lumoral® use and return the remaining Lumorinse® tablets at the 3 and 6-month follow-up visits to determine total consumption. The percentage of compliance was calculated by dividing the number of tablets used by the number of treatment days. They were also asked to observe and self-report any adverse effects of Lumoral® treatment.

### Statistical analysis

GraphPad software version 9.1.0 (GraphPad Software, La Jolla, CA, USA) was used to analyze the data and generate the graphs. Paired T-test and ANOVA were used to compare differences between different time points within a study group. An unpaired T-test was used to compare time points between the groups. A p-value less than 0.05 was considered statistically significant.

## Results

### 3.1. Demographic characteristics of the patient population

Out of 41 Stage I-III randomized patients, 40 were included in the study. One subject was rejected after signing the informed consent before any examinations due to a screening failure. A total of six subjects withdrew from the study before completing their follow-up visits. Five subjects (three in the NSPT + Lumoral® group and two in the NSPT group) withdrew before the six-month follow-up, while one subject in the NSPT + Lumoral® group withdrew before the three-month visit. The study was discontinued for three subjects (two in the NSPT + Lumoral® group and one in the NSPT group) due to unanticipated organizational changes at one of the study centers. The primary researcher at this site transitioned to another position, and no replacement was available to continue the study. Additionally, two subjects in the NSPT + Lumoral® group were diagnosed with a condition unrelated to the study and chose to withdraw voluntarily. Furthermore, one subject in the NSPT + Lumoral® group canceled their final visit and could not be reached for further follow-up. Patient demographics are shown in Table 1.

**Table 1.** Patient demographics.

|  | Lumoral®+NSPT group | NSPT group | Statistical difference |
| --- | --- | --- | --- |
| Gender M/F | 4/17 | 9/11 | p=ns. |
| Age Median (range) | 53 (32-70) | 58.5 (34-69) | p=ns. |
| Chronic Diseases diagnosed Y/N | 16/5 | 15/5 | p=ns. |
| Medication Y/N | 14/7 | 14/6 | p=ns. |
| Diabetes type 2 Y/N | 0/20 | 2/18 | p=ns. |
| Hypertension Y/N | 9/12 | 7/13 | p=ns. |
| Hypercholesterolemia Y/N | 5/16 | 3/17 | p=ns. |
| Asthma Y/N | 1/20 | 3/17 | p=ns. |
| Sleep Apnea Y/N | 3/18 | 0/20 | p=ns. |
| Hypothyroidism Y/N | 2/19 | 0/20 | p=ns. |
| Chronic Bowel Disease | 2/19 | 0/20 | p=ns. |
| Other chronic diagnoses | Psoriasis, Oral Lichen Planus | Meniere, St post chemotherapy | p=ns. |

Table 1. Patient demographics.
|  | **Lumoral®+NSPT**<br>**group** | **NSPT group** | **Statistical**<br>**diffence** |
| --- | --- | --- | --- |
| **Gender M/F** | 4/17 | 9/11 | p=ns. |
| **Age Median (range)** | 53 (32-70) | 58.5 (34-69) | p=ns. |
| **Chronic Diseases diagnosed**<br>**Y/N** | 16/5 | 15/5 | p=ns. |
| **Medication Y/N** | 14/7 | 14/6 | p=ns. |
| **Diabetes type 2 Y/N** | 0/20 | 2/18 | p=ns. |
| **Hypertension Y/N** | 9/12 | 7/13 | p=ns. |
| **Hypercholesterolemia Y/N** | 5/16 | 3/17 | p=ns. |
| **Asthma Y/N** | 1/20 | 3/17 | p=ns. |
| **Sleep Apnea Y/N** | 3/18 | 0/20 | p=ns. |
| **Hypothyroidism Y/N** | 2/19 | 0/20 | p=ns. |
| **Chronic Bowel Disease** | 2/19 | 0/20 | p=ns. |
| **Other chronic diagnoses** | Psoriasis, Oral Lichen<br>Planus | Meniere, St post<br>chemotherapy | p=ns. |

### 3.2. Bleeding on probing

At the baseline, the mean±SEM BOP was 40.2±3,4% in the NSPT + Lumoral® group and 35.5±3.7% in the NSPT group, with no statistical difference between the groups. Both groups showed a reduction in BOP both at the three-month and six-month visits when compared to the baseline, the BOP being 27.0±2.0% and 22.0±1.6%, respectively in the NSPT + Lumoral® group, and 28.2±2.7% and 26.5±2.4%, respectively, in the NSPT group. The BOP reduction was significantly greater in the NSPT + Lumoral® group compared to the NSPT group between the baseline and the final six-month visit (p=0.03). See Figures 2 and 3.

**Figure 2.**
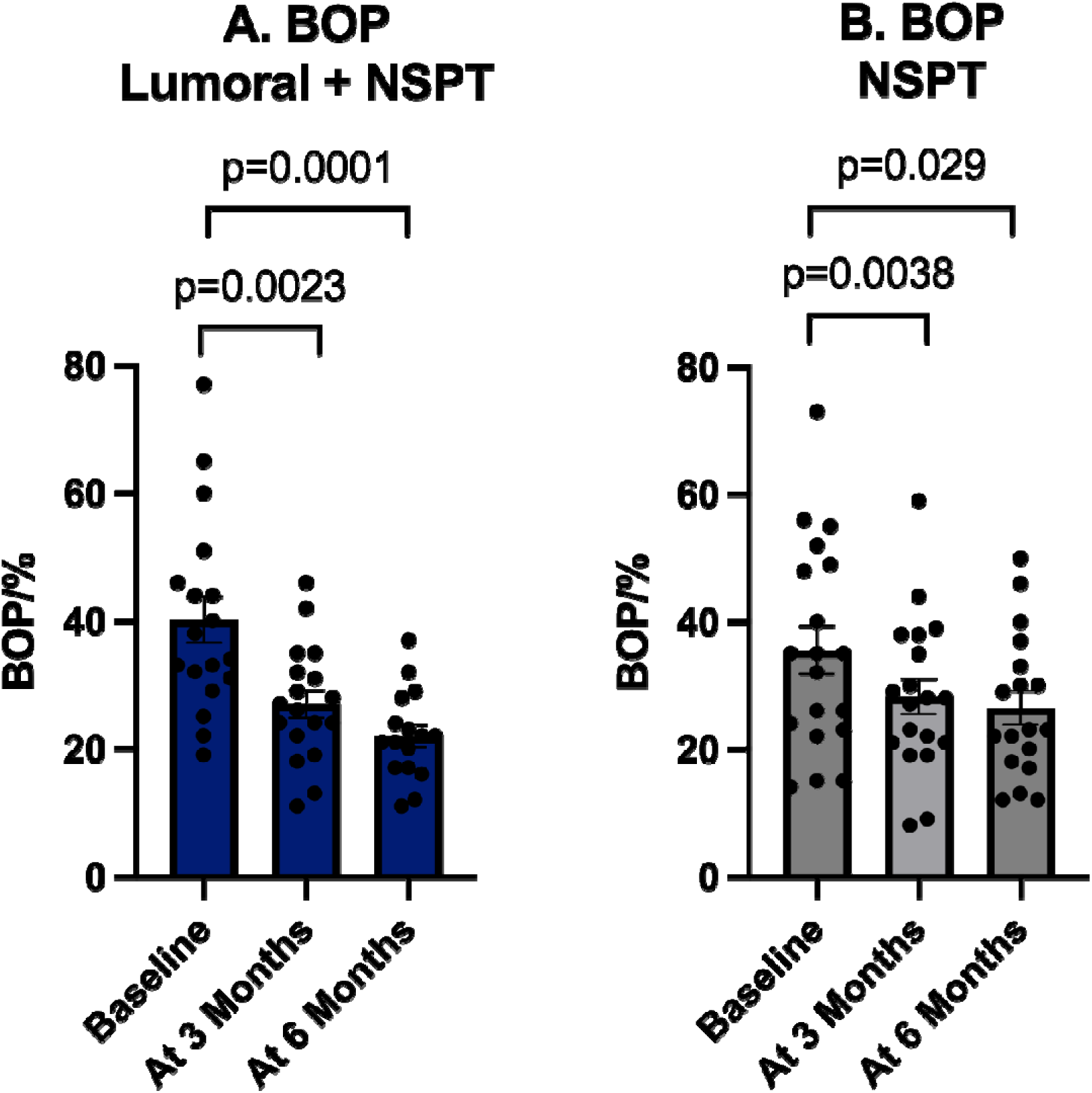
Bleeding on probing improved significantly in both groups.

**Figure 3.**
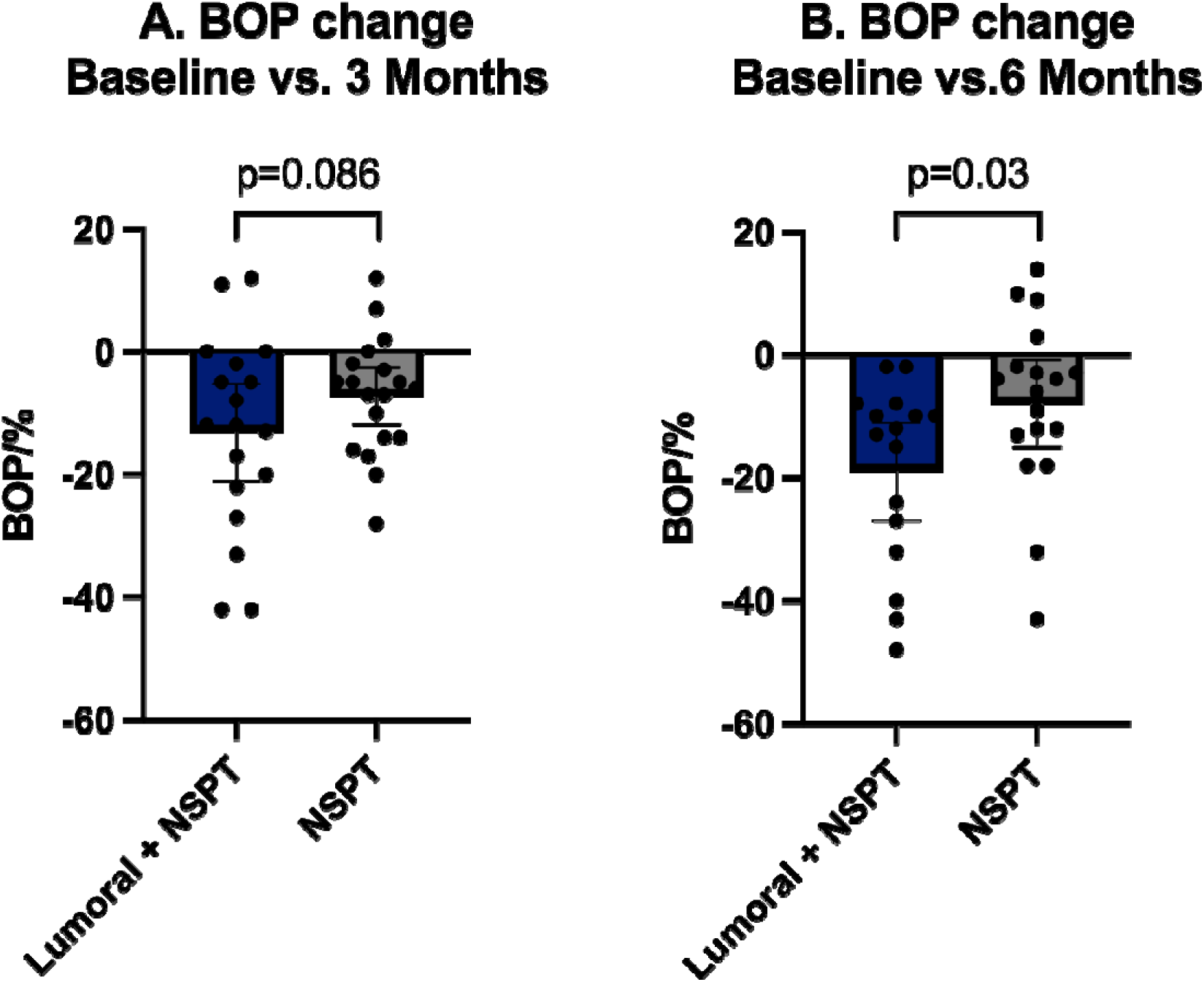
Reduction in the bleeding on probing was greater in the Lumoral® + NSPT group than in the NSPT group.

### 3.2. Plaque index

At the baseline, the mean±SEM VPI was 20.9±4.3% in the NSPT + Lumoral® group and 19.6±3.2% in the NSPT group, with no statistical difference between the groups. At the three-month and six-month visits, the VPI was 14.3±2.4% and 10.6±1.8%, respectively, in the NSPT + Lumoral® group, and 15.0±2.7% and 15.7±4.3%, respectively, in the NSPT group. The NSPT + Lumoral® group had significantly less plaque in the six-month visit compared to the baseline (p=0.04). There was no statistical reduction in the VPI in the NSPT group. See Figure 4.

**Figure 4.**
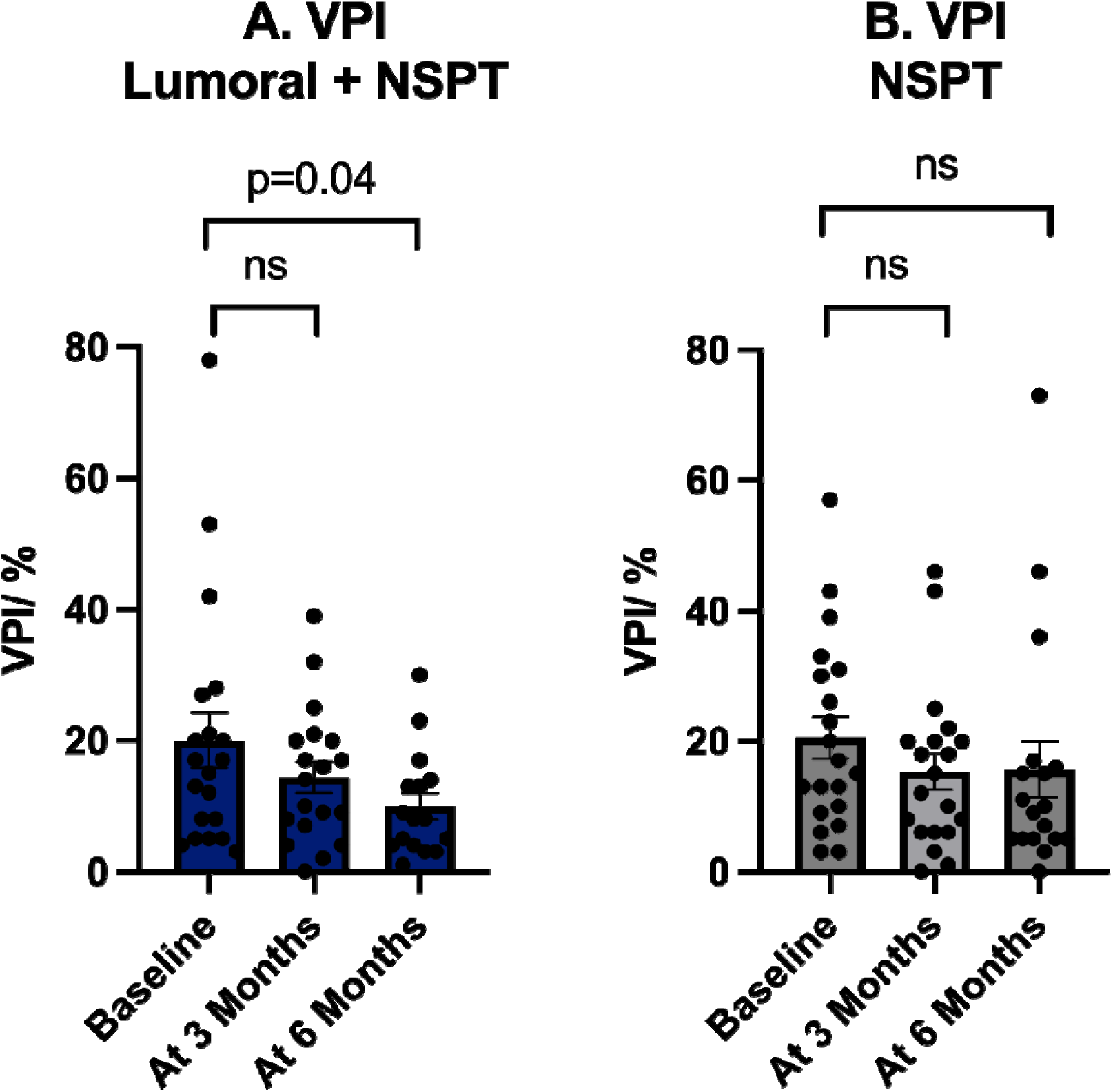
Visual plaque index (%) reduced significantly in the NSPT + Lumoral® group, when no statistical change was observed in the NSPT group.

### 3.3. Probing Pocket Dephts

At the baseline, the number of sites PPD ≥ 4 mm, expressed as mean±sem, was 32.3±4.5. in the NSPT + Lumoral® group and 26.9±3.8 in the NSPT group, with no statistical difference between the groups. In the NSPT + Lumoral® group, at the three-month visit, the number of PPD ≥ 4 mm had significantly reduced down to 24.5±3.7 (p=0.019), and at the six-month visit to 23.7±3.9 (p=0.0043, compared to the baseline). The number of PPD ≥ 4 mm in the NSPT group was 23.8±4.3 and 24.0±4.2 at three-month and six-month visits, respectively. There was no statistically significant reduction in the number of PPD ≥ 4 mm in the NSPT group. See Figure 5.

**Figure 5.**
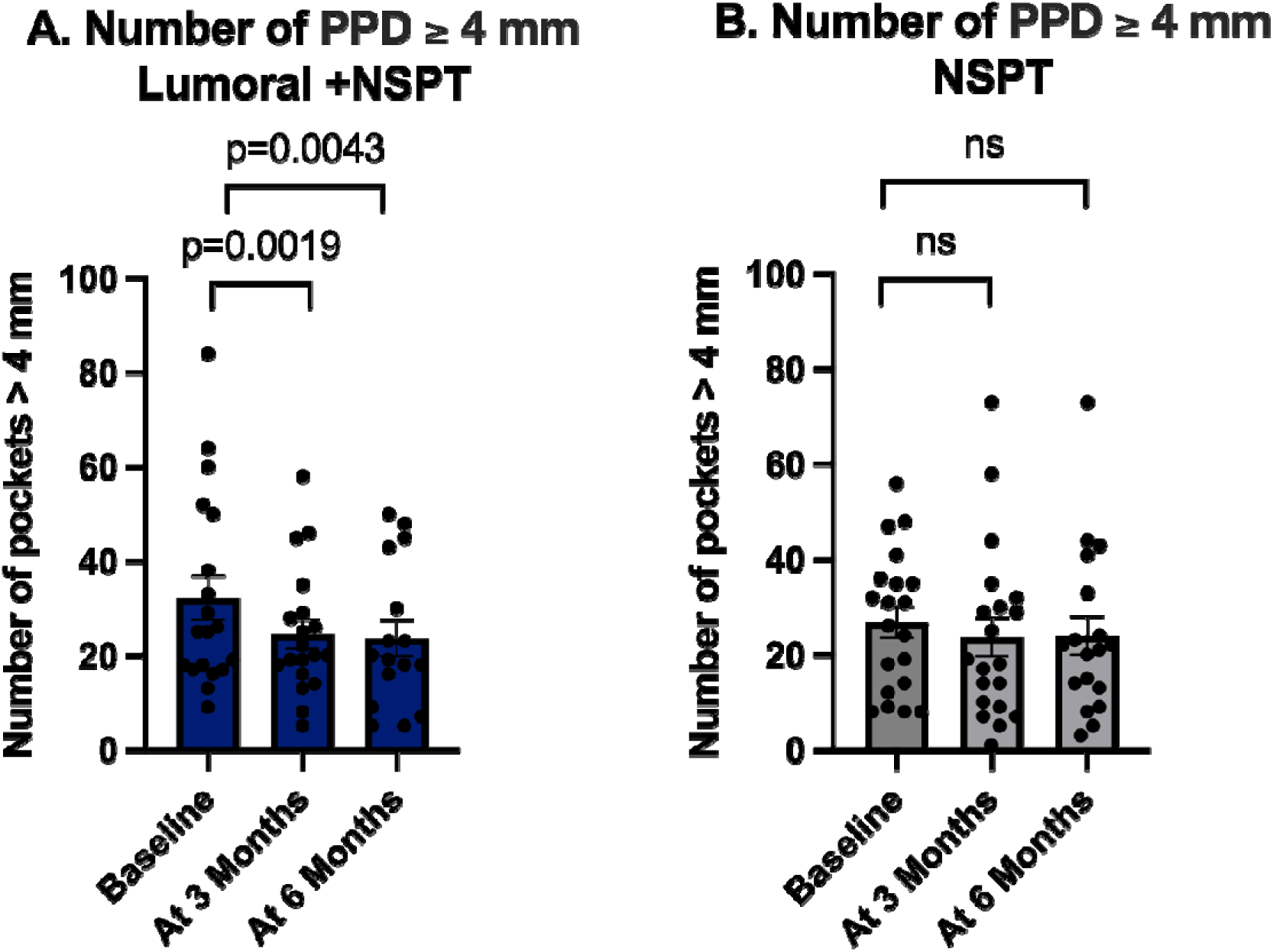
The number of sites with ≥ 4 mm probing pocket depth) reduced significantly in the Lumoral® + NSPT group, while in the NSPT group no statistical change was observed.

### 3.4. PESA and PISA

At the baseline, the PESA, expressed as mean±sem, was 587*±22* mm^2^ in the NSPT + Lumoral® group and 444±16 mm^2^ in the NSPT group, with no statistical difference between the groups. In the NSPT + Lumoral® group, at the three-month visit, the number of deep pockets had significantly reduced, resulting in a significant reduction in the total periodontal epithelial surface area to 451±18 mm^2^ (p=0.017), and at the six-month visit to 414 ±18 mm^2^ (p=0.009, compared to the baseline). The total periodontal epithelial surface area in the NSPT group was 375±18 mm^2^ and 404±17 mm^2^ at three-month and six-month visits, respectively, resulting in no statistically significant reduction of the total periodontal epithelial surface area in the NSPT group. See Figure 6.

**Figure 6.**
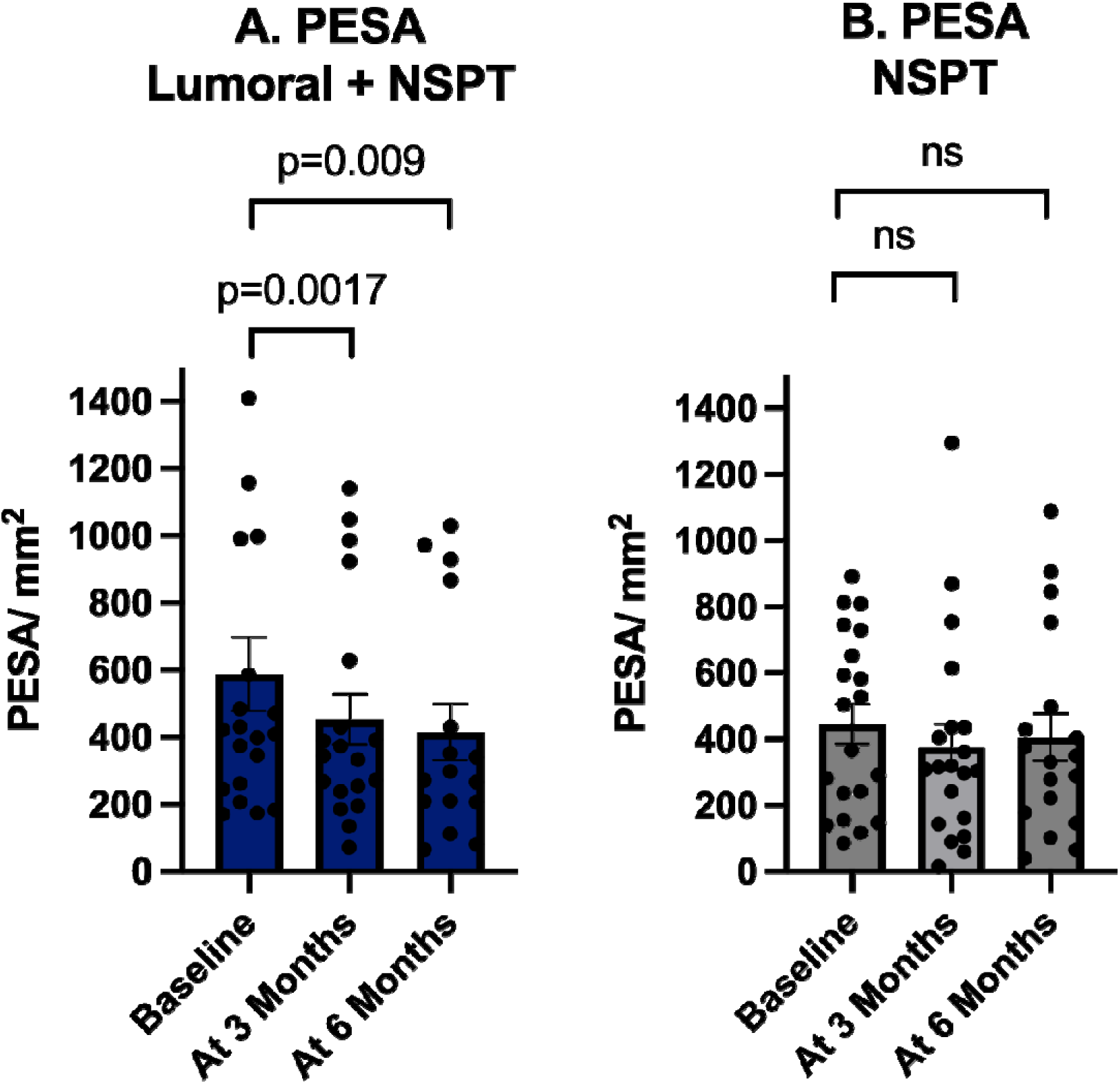
Periodontal epithelial surface area reduced significantly in the Lumoral® + NSPT group during the treatment period, while in the NSPT group, no statistical change was observed.

At the baseline the PISA, expressed as mean±sem, was 385±20 mm^2^. in the NSPT + Lumoral® group and 226±13 mm^2^ in the NSPT group, with no statistical difference between the groups. In the NSPT + Lumoral® group, at the three-month visit, the total periodontal inflamed surface area had significantly reduced to 231±15 mm^2^ (p=0.015), and at the six-month visit to 191±14 mm^2^ (p=0.034, compared to the baseline). The total periodontal inflamed surface area in the NSPT group was 189±14 mm^2^ and 174±13 mm^2^ at three-month and six-month visits, respectively. There was no statistically significant reduction in the number of periodontal pockets in the NSPT group. See Figure 7.

**Figure 7.**
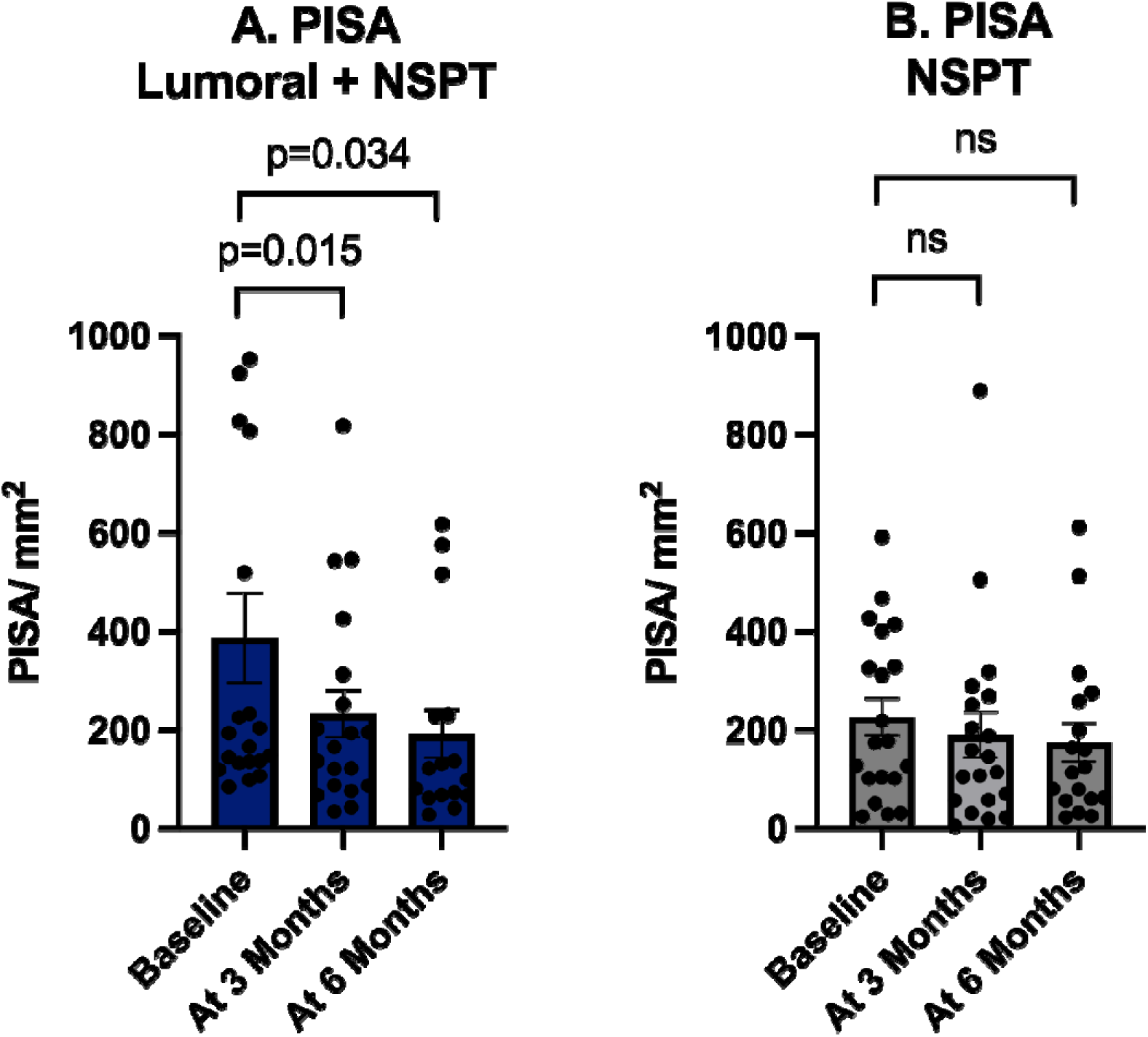
Periodontal inflamed surface area reduced significantly in the Lumoral® + NSPT group, while in the NSPT group no statistical change was observed.

## Discussion

This randomized controlled multicenter study evaluated the clinical benefits and outcomes of adjunctive antibacterial dual-light therapy in the treatment of chronic periodontitis. The Lumoral® device, delivering dual-light antibacterial photodynamic therapy (aPDT) and antibacterial blue light (aBL), was applied regularly at home alongside non-surgical periodontal therapy (NSPT). Both groups received standardized oral hygiene interventions, including home care guidance and electric toothbrushes.

The primary outcome measure was reduction in BOP. Both groups decreased BOP, but the reduction was clearly greater in the NSPT Lumoral® group compared to NSPT alone. What comes to the secondary outcomes, both groups showed improvements in oral hygiene, but plaque reduction was more pronounced in the Lumoral® group. Furthermore, the adjunctive Lumoral® treatment reduced the total number of sites with ≥ 4 mm probing depht. This was also observed, when the total burden of periodontal pockets was measured by PESA, while the NSPT group revealed no significant change. The PESA reduction in the Lumoral® group was observed at both three-months’ and six-months’ visits. Finally, a significant reduction in PISA was observed in the Lumoral® group, with improvements sustained at the six-month follow-up. In contrast, PISA reduction in the NSPT group was not statistically significant.

These findings align with previous studies and further extend them by demonstrating the clear benefits of regular dual-light therapy, including reductions in active matrix metalloproteinase-8 (aMMP-8) levels, BOP, and visible plaque index (VPI) in periodontal patients following intensive Lumoral® use (up to twice daily) [9,16]. Similarly, a recent case report documented significant periodontal health improvements in a patient with impaired oral hygiene ability following regular Lumoral® use [15].

We used PESA and PISA in the assessment of periodontal disease. These are valuable metrics for assessing the overall burden of periodontitis. PESA represents the total surface area of the periodontal epithelium that is exposed due to pocket formation, while PISA quantifies the inflamed portion of that area, reflecting the active disease state. The amount of PESA and PISA are calculated by using the traditional clinical parameters PPD and BOP, which assess localized disease severity. PESA and PISA provide a more comprehensive evaluation of periodontitis by integrating both the extent, severity, and activity of inflammatory burden across the dentition. PISA, in particular, is useful for assessing systemic pro-inflammatory load, often interpreted as periodontal inflammation has been linked to various systemic conditions, including cardiovascular disease and diabetes [19]. By measuring changes in PESA and PISA over time, clinicians can more effectively evaluate treatment responses and monitor disease progression or resolution, making these parameters important tools in both research and clinical practice.

The European Federation of Periodontology (EFP) S3 Level Clinical Practice Guideline (CPG) for Stage I–III periodontitis [20] recommends a stepwise approach to therapy, beginning with behavioral interventions for biofilm control, followed by supra- and subgingival instrumentation, and, if necessary, surgical and supportive periodontal care. The present study aligns with this framework. Both groups received oral hygiene instructions, while additional professional plaque removal in the clinic further contributed to microbial burden reduction. Most participants were in maintenance therapy. Despite adequate disease control, the significant improvements observed with adjunctive Lumoral® therapy emphasize its clear clinical efficiency as an additional tool for periodontitis management.

The antibacterial effect of aPDT relies on activating an externally applied photosensitizer with visible light, generating reactive oxygen species (ROS) that indiscriminately kill bacteria through oxidative stress [2]. Conversely, aBL relies on endogenous bacterial chromophores (e.g., porphyrins and flavins) as internal photosensitizers [3,4]. In fact, studies have demonstrated that the simultaneous application of aPDT and aBL significantly enhances antibactericidal efficacy compared to aPDT alone [12–14,21].

The direct antibacterial effect of adjunctive Lumoral® treatment within periodontal pockets remains uncertain. While 810 nm near-infrared (NIR) light penetrates tissues effectively, the retention of indocyanine green (ICG) in periodontal pockets after rinsing is unclear. Therefore, dual-light therapy should be considered primarily as an enhancement of supragingival plaque control at home, and in a regualar basis,while the subgingival plaque removal should be performed in a clinical setting.

ICG has been widely used in dentistry due to its low toxicity, non-ionizing nature, water solubility, and strong absorption at NIR wavelengths, enabling deep tissue penetration [12,22]. Several studies confirm its efficacy as an adjunctive periodontal therapy. Moro et al. found ICG-based aPDT to be superior to other aPDT approaches [23]. Similarly, a systematic review by Bashir et al. reported an additional mean pocket depth reduction of 1.17 mm at three months and 1.06 mm at six months compared to scaling and root planing (SRP) alone [24].

A major challenge in adjunctive aPDT studies is the heterogeneity of treatment protocols and, more critically, the low frequency of treatment application [25]. A more frequent aPDT administration is challenging when requiring repeated clinical visits, specialized equipment, and trained personnel. Fortunately, advancements in light-emitting diode (LED) technology have enabled the development of personal, handy and practical home-based light therapy devices. Home-based self-administered aPDT allows for more frequent applications, potentially enhancing clinical outcomes. The long-term effects of repetitive aPDT remain poorly studied, making this research a valuable contribution.

NSPT reduces the dysbiotic bacterial burden and inflammation, and promotes periodontal healing. However, mechanical debridement can traumatize inflamed tissues, delaying the healing process. Dual-light therapy with Lumoral® not only provides antimicrobial action but may also enhance tissue healing through photobiomodulation therapy (PBMT). PBMT is a biological process in which photon energy is absorbed by tissue chromophores, mainly cytochrome c oxidase, leading to increased ATP production, DNA/RNA synthesis, nitric oxide release, and cell membrane modulation [26]. PBMT is widely used in oral mucositis treatment, with international guidelines recognizing its effectiveness [27]. A meta-analysis by Peng et al. identified PBMT as the most effective therapy for oral mucositis, with a 95.8% probability of success [28]. However, its efficacy in periodontitis remains controversial, possibly due to low treatment frequencies in published studies. The systematic review by Dalvi et al. found that only two studies applied PBMT at a frequency of 5–10 sessions [26]. While regular Lumoral® therapy theoretically enables frequent PBMT, its precise role in periodontal healing remains unclear, warranting further investigation.

## Conclusion

This multicenter study provides additional evidence that home-based dual-light therapy with Lumoral® may support periodontal health by improving oral hygiene, BOP, and PPD when used as an adjunct to NSPT. The findings highlight the importance of frequent and self-administered therapy, particularly while awaiting the combined effects of aPDT/aBL and the potential benefits of PBMT. Further long-term studies are needed to gain deeper insights into the sustained effects of dual-light therapy in periodontitis management.

## Data Availability

All data produced in the present study are available upon reasonable request to the authors

